# Folate receptor alpha protein expression in ovarian serous cystadenocarcinoma tumors of The Cancer Genome Atlas: exploration beyond single-agent therapy

**DOI:** 10.1101/2024.04.12.24305742

**Authors:** Christianne Persenaire, Benjamin G. Bitler, Bradley R. Corr

## Abstract

Epithelial ovarian cancer (EOC) can be highly lethal, with limited therapeutic options for patients with non-homologous recombination deficient (HRD) disease. Folate receptor alpha (FOLR1/FRα)-targeting agents have shown promise both alone and in combination with available therapies, but the relationship of FRα to other treatment-driving biomarkers is unknown. The Cancer Genome Atlas (TCGA) was queried to assess protein and mRNA expression and mutational burden in patients with differential FRα protein-expressing ovarian tumors, and the results referenced against the standard 324 mutations currently tested through FoundationOne Companion Diagnostics to identify targets of interest. Of 585 samples within TCGA, 121 patients with serous ovarian tumors for whom FRα protein expression was quantified were identified. FRα protein expression significantly correlated with FOLR1 mRNA expression (p=7.19×10^-14^). Progression free survival (PFS) for the FRα-high group (Q1) was 20.7 months, compared to 16.6 months for the FRα-low group (Q4, Logrank, p=0.886). Overall survival (OS) was 54.1 months versus 36.3 months, respectively; however, this result was not significant (Q1 vs. Q4, Logrank, p=0.200). Mutations more commonly encountered in patients with high FRα-expressing tumors included PIK3CA and FGF family proteins. Combinations of FRα-targeting agents with PI3K, mTOR, FGF(R) and VEGF inhibitors warrant investigation to evaluate their therapeutic potential.

**Simple Summary:** Epithelial ovarian cancer can be highly lethal, with limited therapeutic options for patients without BRCA mutations or non-homologous recombination deficient disease. Folate receptor alpha (FRα)-targeting agents have shown promise in the setting of platinum-sensitive and platinum-resistant ovarian cancer, both alone and in combination with available therapies, but the relationship of FRα to other treatment-driving biomarkers is unknown. This study identifies potential targetable mutations in FRα-expressing tumors, including PIK3CA and FGF/R family proteins, and provides a basis for future investigations of novel combinations of FRα-targeting agents with PIK3CA, mTOR, FGF/R, and VEGF inhibitors.

## 1. Introduction

Ovarian cancer is the second most common gynecologic malignancy affecting women in the United States and accounts for more deaths than all other gynecologic malignancies combined. The standard of care for women with epithelial ovarian cancer (EOC), including ovarian, fallopian tube, and primary peritoneal cancers, remains a combination of cytoreductive surgery and taxane/platinum-based chemotherapies. However, despite advancements in treatment with approved targeted therapies such as poly(ADP-ribose) polymerase (PARP) inhibitors and vascular endothelial growth factor (VEGF) inhibitors, disease recurrence still occurs at an alarming rate of 70-80% [1]. Recurrent disease most commonly progresses to a non-curable, platinum-resistant state, and there remains a critical need to develop new therapeutic approaches.

Over the past decade, folate and its membrane-bound receptor have garnered increased interest as a potential molecular target, as rapidly dividing cancer cells are highly dependent on folate metabolism for DNA replication [2]. Folate receptor alpha (FOLR1/FRα), specifically, is a transmembrane protein involved in cellular folate transport whose expression is highly restricted in somatic tissues. Approximately 80% of EOC constitutively overexpress this receptor, and elevated expression has been found to be associated with a more aggressive tumor phenotype [3].

FRα-targeting agents range in design from monoclonal antibodies and small molecules to folate-drug and antibody-drug conjugates (ADCs). Farletuzumab, for example, is a fully humanized IgG1 antibody that targets FRα-positive cancer cells, while CT900 is a small molecule thymidylate synthase inhibitor that is transported into tumor cells by its high affinity for the FRα receptor [4]. Vintafolide is a folate-drug conjugate in which a water-soluble folic acid derivative is linked through a peptide spacer to a potent microtubule destabilizing agent called desacetylvinblastine monohydrazide, a vinca alkaloid [4]. Once released within the cell, the vinca alkaloid acts to inhibit the beta-tubulin polymerization necessary for cell division to induce cell cycle arrest.

ADCs are a novel class of antineoplastic agents that can utilize FRα as a biomarker-driven target. These entities typically consist of a monoclonal antibody directed against a tumor-associated antigen to which a cytotoxic agent (‘payload’) is conjugated; once internalized, the payload is released to mediate its cytotoxic effects. This design has the theoretical benefit of minimizing systemic toxicity while maximizing efficacy [5], and the inherent capacity of FRα to internalize large molecules makes the receptor ideally suited for targeting with ADCs such as mirvetuximab soravtansine (MIRV), STRO-002, and MORab-202. Along with farletuzumab, these ADCs, as well as ELU001, a C’Dot drug conjugate (NCT05001282); TPIV200/hu-FR-1, a multi-epitope anti-folate receptor vaccine (NCT02764333) [6]; and ITL-306, a cell-based therapy derived from tumor-infiltrating lymphocytes (TIL) whose activity increases when the agent encounters tumor-specific FRα (NCT05397093); are under active investigation.

MIRV is an ADC with the most clinical evaluation in EOC of its drug class to date. It is comprised of an FRα-binding antibody, cleavable linker, and the maytansinoid DM4, a potent tubulin-targeting agent. Upon antigen binding, the FRα-ADC complex is rapidly internalized and DM4 is released to induce cell cycle arrest and apoptosis. MIRV has the additional benefit of bystander killing, as the cleavable linker design allows active DM4 metabolites to diffuse from the antigen-positive tumor cells into neighboring cells [7]. STRO-002 is another FRα-specific ADC comprised of an anti-FRα human IgG1 antibody, SP8166, conjugated to a cleavable DBCO-3-aminophenyl-hemiasterlin drug-linker, while MORab-202 is an ADC in which farletuzumab and eribulin, a microtubule inhibitor, are linked via cleavable cathepsin B [8].

Early clinical studies of MIRV in patients with platinum-resistant ovarian cancer (PROC) showed promise, with an objective response rate (ORR) of 39% in a phase Ib trial with few grade 3/4 toxicities [9]. In November of 2022, MIRV was granted accelerated approval for FRα positive platinum-resistant ovarian cancer based on results from the SORAYA trial (NCT04296890). Single-agent use in patients with up to three prior lines of therapy, including required bevacizumab, demonstrated an ORR of 32.4%, including five complete responses (CR), and a median duration of response (DOR) of seven months. Response rates were similar regardless of the number of prior therapies received, including PARP inhibitors [10]. More recently, encouraging results from the international phase III randomized controlled trial, MIRASOL, comparing MIRV to investigator’s choice chemotherapy (IC) in patients with FRα positive PROC, were presented at the 2023 Annual Meeting of the American Society of Clinical Oncology (ASCO). Study authors found the median overall survival (OS) was 16.5 months with MIRV vs. 12.8 months with IC (HR 0.67, p = 0.0046), while the median progression free survival (PFS) was 5.62 months vs 3.98 months in each respective arm (HR 0.65, p <0.0001). Objective response rate was 42.3% in the MIRV group versus 15.9% in the IC group (p <0.0001). These findings made MIRV the first treatment to demonstrate a PFS and OS benefit in PROC compared to IC [11].

Other ADCs have had similarly encouraging results. In heavily pretreated patients with advanced EOC, for example, STRO-002 had an ORR of 33% in 33 evaluable patients across all FRα expression levels [12]. A phase I study of MORAb-202 in 22 patients showed a CR, partial response (PR), and stable disease in one, nine, and eight patients, respectively [13].

Prior efforts to target FRα with farletuzumab and vintafolide were hampered by limited single-agent activity [14, 15]. However, vintafolide showed a statistically significant improvement when used in combination with pegylated liposomal doxorubicin [16], suggesting FRα-targeting agents may prove more effective when used in combination with other agents [17]. Combinatory strategies with other FRα targeting agents are currently under investigation. For example, clinical trials with MIRV in combination with carboplatin (NCT04606914), rucaparib (NCT03552471), bevacizumab (NCT05445778), pegylated liposomal doxorubicin or pembrolizumab (NCT02606305), and gemcitabine (NCT02996825) are underway. Similarly, a study combining STRO-002 with bevacizumab (NCT05200364) is actively recruiting.

As the relationship of FRα to other treatment-driving biomarkers is currently unknown, the objective of this study was to analyze TCGA to assess protein/mRNA expression and mutational burden in patients with differential FRα protein-expressing tumors and identify potential targets for combination drug therapies.

## 2. Materials & Methods

The Cancer Genome Atlas (TCGA) Ovarian Serous Cystadenocarcinoma PanCancer Atlas was accessed via cBioportal (http://www.cbioportal.org) and queried in May 2022 to generate a list of patients for whom data regarding tumor levels of FRα protein expression were available. Protein expression was calculated based on the Clinical Proteomic Tumor Analysis Consortium (CPTAC, Figure 1A) [18].

**Figure 1:**
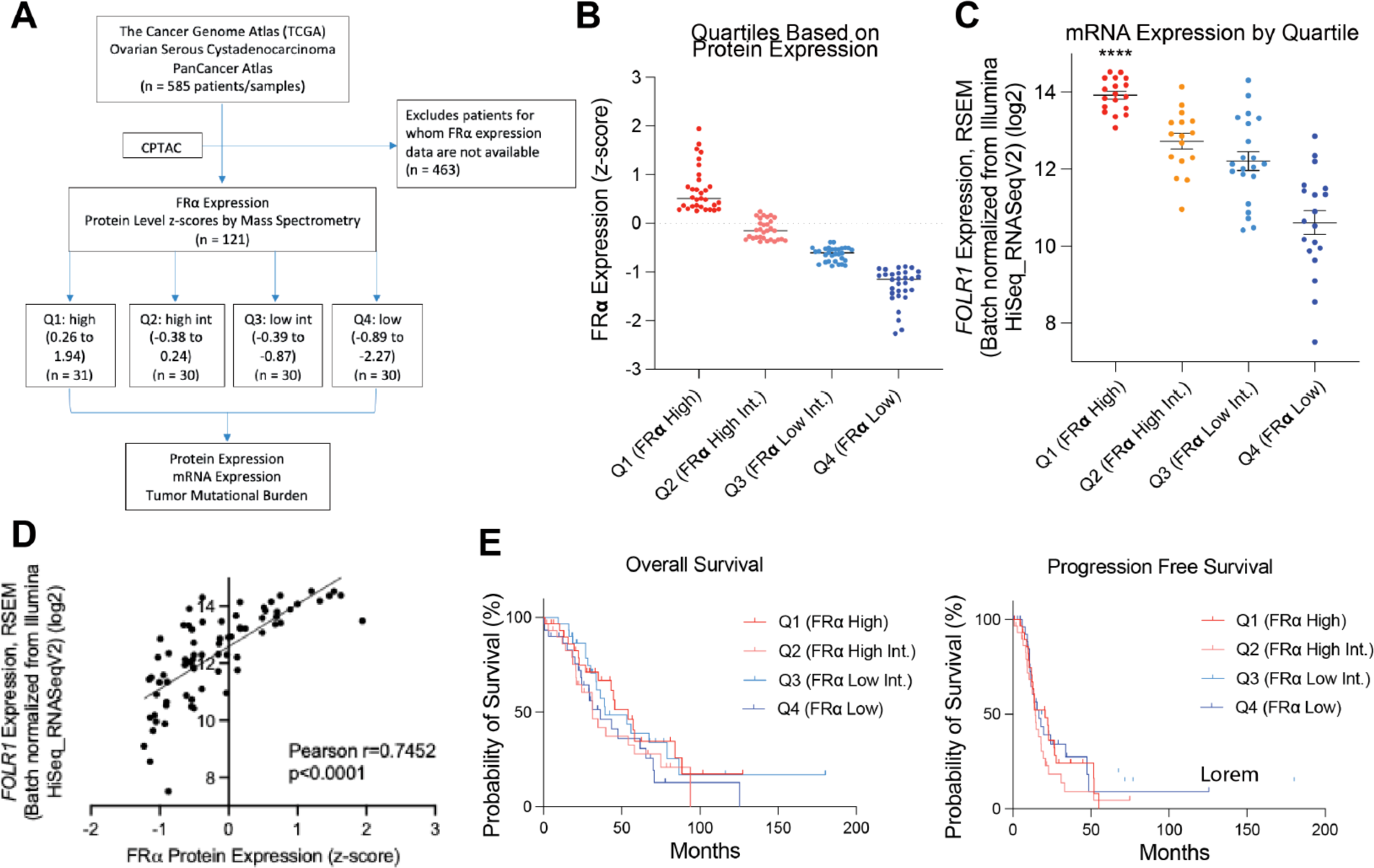
(A) Consort diagram establishing the TCGA PanCancer Atlas patient population for whom FRα expression data were available. (B) Columnar scatter plot of FRα protein expression values by quartile. (C) Columnar scatter plot of FRα mRNA expression by quartile. (D) Scatter plot depicting Pearson correlation of 0.75 between FOLR1 mRNA transcription and FRα protein expression. (E) Kaplan Meier curves depicting overall survival (OS) and progression free survival (PFS) by quartile.

We stratified patients into quartiles based on FRα expression, where Q1 = FRα high, Q2 = FRα high-intermediate (int.), Q3 = FRα low-int., and Q4 = FRα low. These quartiles were used to perform survival analyses and protein expression (Z-score) analyses, as well as to examine mRNA expression and gene mutations as a function of FRα expression. The results were then referenced against the standard 324 mutations currently tested through FoundationOne Companion Diagnostics (CDx), including those responsible for microsatellite instability, tumor mutational burden and loss of heterozygosity, to identify targets of interest. Statistical analyses were conducted and graphs generated in Prism GraphPad (v9.0) using Kaplan Meier curves, as well as Logrank, Chi-squared, Spearman’s rho, Pearson’s rho, Benjamini-Hochberg and student t-tests.

## 3. Results

Utilizing TCGA PanCancer Atlas (n=585), we identified 121 patients with serous ovarian tumors for whom FRα protein expression was quantified. Those patients were then stratified into quartiles to assess for correlation with outcomes of interest (Figure 1A-B). TCGA PanCancer Atlas comparison of mRNA expression, protein expression (CPTAC) and genomic alterations was utilized across the four quartiles and data collected for 18,695 mRNA transcripts and 7,620 proteins. Both mRNA expression and protein expression of FRα decreased in stepwise fashion from Q1 to Q4 (Figure 1B-C), and FOLR1 mRNA expression was significantly elevated (t-test; p=7.19×10^-14^; FDR q=6.78×10^-10^) across the different quartiles (Figure 1C). The Pearson correlation between FOLR1 mRNA transcription and FRα protein expression was 0.7452 (p<0.0001, Figure 1D).

When evaluating survival outcomes by quartile, PFS for the FRα-high group (Q1) was 20.7 months, compared to 16.6 months for the FRα-low group (Q1 vs Q4, Logrank, p=0.886). Overall survival was 54.1 months versus 36.3 months, respectively; however, this result was not significant (Q1 vs. Q4, Logrank, p=0.200) (Figure 1E, Table 1).

**Table 1:** Overall survival and progression free survival in patients with EOC stratified by levels of tumor FRα expression; p-values derived by Log Rank test.

|  | Median Survival in Months |  |  |  |  |
| --- | --- | --- | --- | --- | --- |
|  | Q1 (High FOLR1) | Q2 (High Int. FOLR1) | Q3 (Low Int FOLR1) | Q4 (Low FOLR1) | p-value |
| Overall Survival | 54.1 | 31.3 | 39.4 | 36.3 | 0.3 |
| Months (95% CI) | (43.2-NA) | (20.9-74.9) | (33.9-NA) | (24.2-70.1) |  |
| Progression Free Survival | 20.7 | 14.5 | 16 | 16.6 | 0.4 |
| Months (95% CI) | (12.9-27.8) | (11.8-19.6) | (13.0-26.9) | (11.54-NA) |  |

To identify potential molecular targets correlating to FRα expression, data for the 324 genes assessed through FoundationOne CDx were collected, as these represent mutations more frequently seen in patients with solid tumors, including EOC, and many represent areas of active investigation for which targeted therapies exist, or are currently in development. From these data, we compiled a list of the top thirty mutations across the FRα quartiles, with results as noted in Table 2. Spearman’s rank correlation coefficient was calculated for protein expression for genes part of FoundationOne CDx to identify proteins whose expression was strongly positively or negatively associated with that of FRα protein; these results are shown in Table 3.

**Table 2:** Most common mutations encountered in tumors with high, high-intermediate, low-intermediate, and low FRα expression; p-values derived by Chi squared test.

| Gene Symbol | Number of mutations (%) |  |  |  | p-value |
| --- | --- | --- | --- | --- | --- |
|  | Q1 (High FOLR1) | Q2 (High Int. FOLR1) | Q3 (Low Int FOLR1) | Q4 (Low FOLR1) |  |
| PIK3CA | 9 (29.03%) | 5 (16.67%) | 7 (23.33%) | 6 (20.69%) | 0.703 |
| ETV5 | 7 (22.58%) | 6 (20.00%) | 4 (13.33%) | 4 (13.79%) | 0.724 |
| FGF12 | 7 (22.58%) | 7 (23.33%) | 6 (20.00%) | 4 (13.79%) | 0.793 |
| KLHL6 | 7 (22.58%) | 6 (20.00%) | 4 (13.33%) | 6 (20.69%) | 0.814 |
| RAD21 | 7 (22.58%) | 9 (30.00%) | 11 (36.67%) | 7 (24.14%) | 0.61 |
| CCND1 | 6 (19.35%) | 2 (6.67%) | 2 (6.67%) | 0 (0.00%) | 0.0507 |
| FGF19 | 6 (19.35%) | 2 (6.67%) | 2 (6.67%) | 0 (0.00%) | 0.0507 |
| FGF3 | 6 (19.35%) | 2 (6.67%) | 2 (6.67%) | 0 (0.00%) | 0.0507 |
| FGF4 | 6 (19.35%) | 2 (6.67%) | 2 (6.67%) | 0 (0.00%) | 0.0507 |
| MAP3K13 | 6 (19.35%) | 7 (23.33%) | 5 (16.67%) | 4 (13.79%) | 0.807 |
| SOX2 | 6 (19.35%) | 5 (16.67%) | 5 (16.67%) | 6 (20.69%) | 0.971 |
| IRF4 | 5 (16.13%) | 3 (10.00%) | 4 (13.33%) | 1 (3.45%) | 0.431 |
| TIPARP | 5 (16.13%) | 3 (10.00%) | 4 (13.33%) | 3 (10.34%) | 0.876 |
| ERBB4 | 5 (16.13%) | 2 (6.67%) | 1 (3.33%) | 0 (0.00%) | 0.0698 |
| ATR | 5 (16.13%) | 4 (13.33%) | 2 (6.67%) | 1 (3.45%) | 0.332 |
| TERT | 4 (12.90%) | 3 (10.00%) | 2 (6.67%) | 0 (0.00%) | 0.266 |
| BCL6 | 4 (12.90%) | 6 (20.00%) | 4 (13.33%) | 5 (17.24%) | 0.856 |
| RSPO2 | 4 (12.90%) | 8 (26.67%) | 3 (10.00%) | 7 (24.14%) | 0.257 |
| XRCC2 | 4 (12.90%) | 4 (13.33%) | 4 (13.33%) | 1 (3.45%) | 0.539 |
| BRD4 | 4 (12.90%) | 2 (6.67%) | 0 (0.00%) | 6 (20.69%) | 0.0532 |
| NOTCH3 | 4 (12.90%) | 2 (6.67%) | 1 (3.33%) | 6 (20.69%) | 0.149 |
| SDHA | 4 (12.90%) | 3 (10.00%) | 2 (6.67%) | 0 (0.00%) | 0.266 |
| CCNE1 | 4 (12.90%) | 4 (13.33%) | 5 (16.67%) | 7 (24.14%) | 0.632 |
| BRAF | 4 (12.90%) | 5 (16.67%) | 3 (10.00%) | 4 (13.79%) | 0.899 |
| ERBB3 | 4 (12.90%) | 1 (3.33%) | 0 (0.00%) | 1 (3.45%) | 0.113 |
| MCL1 | 4 (12.90%) | 2 (6.67%) | 1 (3.33%) | 2 (6.90%) | 0.552 |
| BRCA1 | 3 (9.68%) | 1 (3.33%) | 1 (3.33%) | 2 (6.90%) | 0.665 |
| BTG2 | 3 (9.68%) | 2 (6.67%) | 0 (0.00%) | 1 (3.45%) | 0.344 |
| CDKN1A | 3 (9.68%) | 3 (10.00%) | 1 (3.33%) | 0 (0.00%) | 0.27 |
| FOXL2 | 3 (9.68%) | 4 (13.33%) | 1 (3.33%) | 1 (3.45%) | 0.373 |

**Table 3:** Proteins with an absolute Spearman’s Rho >0.98 suggesting association with FOLR1 expression; p-values derived by student’s t-test.

| Gene Symbol | Gene Name | FOLR1 Protein Expression (Z-score) |  |  |  | Rho value | p-value |
| --- | --- | --- | --- | --- | --- | --- | --- |
|  |  | Q1 (High) | Q2 (High Int) | Q3 (Low Int) | Q4 (Low) |  |  |
| **CTNNA1** | alpha catenin | 0.14 | 0.03 | -0.02 | -0.07 | 0.991 | 0.002 |
| **FH** | fumarate hydratase | 0.16 | 0 | -0.05 | -0.12 | 0.985 | 0.013 |
| **CDH1** | e-cadherin | 0.09 | 0.01 | -0.08 | -0.21 | 0.984 | 0.008 |
| **SDHA** | succinate dehydrogenase complex flavoprotein subunit A | 0.09 | 0 | -0.01 | -0.07 | 0.981 | 0.110 |
| **SRC** | proto-oncogene C-Src | -0.08 | -0.05 | 0.01 | 0.05 | -0.977 | 0.271 |
| **AKT3** | protein kinase B, gamma | -0.2 | -0.01 | 0.04 | 0.12 | -0.981 | 0.529 |
| **NF1** | neurofibromin 1 | -0.09 | -0.06 | -0.01 | 0.03 | -0.982 | 0.627 |
| **QKI** | quaking homolog, KH domain RNA binding | -0.05 | 0.03 | 0.07 | 0.13 | -0.999 | 0.012 |

Only 160/324 genes on the FoundationOne CDx were identified via CPTAC. Quaking Homolog, KH Domain RNA Binding (QKI) was the most negatively correlated (Pearson r = -0.996) and Alpha Thalassemia/Mental Retardation Syndrome X-Linked (ATRX) was the most positively correlated (Pearson r = 0.993). BRCA1 protein expression was undetectable and BRCA2 protein expression did not correlate with FRα protein expression (Pearson r = 0.27). Data regarding protein expression for other genes in the HRD pathway were available only for ATM, CHEK2 and NBS1, and also showed no correlation.

## 4. Discussion

Targeting FRα is an area of active investigation and one that holds significant promise. In this study, we used TCGA to identify potential molecular targets that could potentiate the ORR of FRα-targeting agents and improve their therapeutic efficacy for patients with EOC, particularly those with non-HRD, platinum-resistant disease. We identified an increased frequency of potentially targetable PIK3CA and FGF family protein mutations in FRα-expressing tumors.

### Results in the Context of Published Literature

Although early studies of single-agent MIRV, STRO-002, and MORAb-202 suggested these have anti-tumor activity, several combinatorial approaches are under active investigation. MIRV, for example, is being studied in combination with various chemotherapies and targeted therapies such as pembrolizumab and bevacizumab (NCT02606305). In 2018, as part of FORWARD II, Moore et al. published data from the phase 1b escalation study of MIRV in combination with carboplatin in patients with platinum-sensitive ovarian cancer (PSOC), which found a 71% ORR, including three CR and nine PR, and a median PFS of 15 months [19]. Among others, these findings prompted a single arm, phase II trial of carboplatin and MIRV for first-line treatment of patients with advanced, FRα-positive EOC receiving neoadjuvant chemotherapy (NCT04606914). A randomized phase II trial of MIRV in FRα-high, recurrent PSOC also began enrolling patients in September 2021, with the goal of comparing platinum-based chemotherapy followed by PARPi (if indicated) to carboplatin + MIRV followed by MIRV monotherapy (NCT04274426).

For patients with PROC, another expansion cohort from FORWARD II found a confirmed ORR of 43% in patients treated with a combination of MIRV and pembrolizumab, with a median DOR of 6.9 months and median PFS of 5.2 months [20]. The same phase Ib study included a cohort of patients with PROC who received MIRV in combination with bevacizumab; for these patients, the confirmed ORR was 39%, including five CR and 21 PR; median PFS was 6.9 months. Patients who were bevacizumab-naïve, less heavily pretreated, and whose tumors had medium/high FRα expression had an ORR of 56% with a median DOR of 12 months and PFS of 9.9 months [21]. Given these findings, FORWARD II looked at the combination of MIRV, carboplatin and bevacizumab followed by MIRV and bevacizumab maintenance therapy in 41 patients with recurrent PSOC, with a confirmed ORR of 83%, median DOR of 10.9 months and PFS of 12.8 months [22]. These data suggest synergistic and durable anti-tumor effects of combinatorial approaches, and more trials are in active development.

In our effort to identify potential targets, we found *PIK3CA* is mutated in 29% of tumors with high FRα expression. PIK3CA, or phosphatidylinositol-4,-5-bisphosphanate 3-kinase catalytic subunit alpha, is an oncogene that has been implicated in several cancers, including breast, colon, liver, and stomach cancers [23]. Point mutations in this gene can cause hyperactivation of the alpha isoform (p110a) of PI3K, which in turn results in constitutive activation of the intracellular PI3K/Akt signaling pathway. This pathway is linked to several critical physiologic and pathophysiologic functions, including those that drive tumor progression, such as cell proliferation, growth, metabolism, angiogenesis, and motility [24]. Several PI3K inhibitors, such as alpelisib, a small molecule, a-specific PI3K inhibitor that selectively inhibits p110a, are being evaluated, and early work suggests that PIK3CA-mutated cancers are sensitive to treatment with these agents [25]. When given in combination with standard of care therapies, as with olaparib in ovarian cancer or fulvestrant in breast, these drugs may work synergistically to overcome tumor resistance and sensitize the tumor cells to treatment with currently approved agents [26, 27].

The propensity toward *PIK3CA* mutations in FRα high tumors also provides a basis for investigating combinations of FRα-targeting agents with mammalian target of rapamycin, or mTOR, inhibitors. This is because activation of the PI3K/Akt signaling pathway results in downstream activation of the mTOR pathway. As with the PI3K/Akt pathway, mTOR plays a key role in regulating cell proliferation, and functions as a regulator of autophagy and apoptosis; thus, alterations in this pathway can lead to unchecked cell proliferation. In ovarian cancer, specifically, the PI3K/Akt/mTOR pathway has been found to activated in approximately 70% of cases and has been linked to higher invasive and migratory capacity. The mTOR inhibitors temsirolimus and everolimus have been studied in clinical trials of patients with recurrent EOC and had promising results [28].

Importantly, while VEGF was not one of the top mutations identified based on FRα expression, receptor binding of VEGFR2 has been found to activate PI3K and Akt. Although the mechanism by which this occurs is not fully understood, it is thought to occur by TSAd-mediated activation of Src family kinases, which engage the receptor tyrosine kinase Axl to trigger ligand-independent autophosphorylation and promote association with PI3K and activation of Akt [29]. This association between VEGFR2 and PI3K/Akt activation, as well as the higher rate of PIK3CA mutations in FRα-high tumors, may provide a mechanistic basis for the promising therapeutic efficacy seen in the early trials of patients treated with combinations of MIRV with bevacizumab mentioned previously. A phase 3 trial to evaluate MIRV in combination with bevacizumab versus bevacizumab alone in PSOC (GLORIOSA) began recruiting patients in December 2022 (NCT 05445778).

Other mutations more frequently observed in FRα-high tumors were those of the fibroblast growth factor, or FGF, family, including FGF3 (19.35% vs. 0%), FGF4 (19.35% vs. 0%), FGF12 (22.58% vs. 13.79%), and FGF19 (19.35% vs. 0%). Broadly speaking, the fibroblast growth factor receptor (FGFR) family of receptor tyrosine kinases consists of four transmembrane receptors, FGFR1-4, and twenty-two known FGF ligands that interact to activate downstream signaling pathways through an intracellular domain. One such pathway is the extracellular signal-regulated kinase (ERK)/mitogen-activated protein kinase (MAPK) pathway, which promotes cell survival, proliferation, angiogenesis, and differentiation—processes exploited in malignant transformation and tumorigenesis [30]. Several agents targeting FGF have been developed for clinical use, ranging from selective tyrosine kinase inhibitors and monoclonal antibodies to ADCs and FGF ligand traps. Two anti-FGFR therapies have been approved by the Food and Drug Administration to date, including erdafitinib for FGFR-3-altered urothelial cancer, and pemigatinib for FGFR-2 fusion cholangiocarcinoma [30]. Lenvatinib, an oral receptor tyrosine kinase inhibitor that has activity against FGFR1-4, as well as VEGFR1-3, platelet derived growth factor receptor-beta, and the RET and kit proto-oncogenes, is currently approved for patients with advanced endometrial cancer without microsatellite instability. Given the higher FGF mutation rate seen in FRα-high tumors in this study, investigation into combinations of FRα-targeting therapies with anti-FGF/R modalities may prove a fruitful area of inquiry.

It should be noted that the BRCA and HRD pathway genes did not demonstrate a clear correlation with FRα expression. The combinatory strategy with PARP inhibitors was first assessed in NCT03552471 utilizing MIRV and rucaparib in recurrent ovarian, fallopian tube, peritoneal or endometrial cancers. The ORR was 29% in the ovarian cancer cohort with a PFS >6months in 47.1% [31]. The lack of correlation of FRα expression and BRCA/HRD demonstrates that a synergistic effect between the two is unlikely, but an additive effect in distinct patient populations may warrant future investigation.

### Strengths and Limitations

A pitfall from previous trials was the method of determining FRα expression in primary tumors, which is critical, as subsequent trials observed a strong positive correlation between FRα expression and ORR. One of the strengths of this analysis was the finding that FRα protein expression significantly correlates with FOLR1 mRNA expression. This raises the possibility of examining mRNA expression in lieu of immunohistochemical based assays in future studies. Limitations of this study include those intrinsic to the use of publicly available datasets such as TCGA PanCancer Atlas, which has restricted scope with a limited number of analyzed samples. In this case, data regarding FRα expression were available for only 121 of 585 patients included in the PanCancer Atlas. An additional limitation is the confounding effect of chromosomal locus on the amplification or overexpression of a target of interest. FOLR1 is found on chromosome 11; the differential expression of proteins found on chromosome 11 may thus be related to proximity to the FRα gene, resulting in similar transcriptional regulation.

## 5. Conclusion

To date, only a few targetable mutations have been examined as part of combinatorial approaches to the treatment of ovarian cancer; based on our findings, there are multiple avenues for further investigation that have the potential to improve outcomes for patients with advanced EOC who currently lack other therapeutic options. In light of the promising results of MIRASOL, this work provides a compelling indication to explore and/or expand upon potential drug combinations of FRα-targeting agents with PI3K, mTOR and FGF/FGFR inhibitors, and bevacizumab to evaluate their therapeutic potential in advanced EOC.

## Data Availability

All data produced in the present study are available upon reasonable request to the offers.

http://www.cbioportal.org

## Author Contributions

Each author was actively involved in this research and approved the final version for submission. The individual contributions are listed below:

Christianne Persenaire: drafted written manuscript, created figures, and contributed to reviews

Benjamin G. Bitler: conceptual design, drafted written manuscript, created figures, and contributed to reviews

Bradley R. Corr: conceptual design, drafted written manuscript, and contributed to reviews

## References

1. Pignata S, C Cecere S, Du Bois A, Harter P, Heitz F. Treatment of recurrent ovarian cancer. Ann Oncol. 2017;28(suppl_8):viii51–viii56. Doi:10.1093/annonc/mdx441

2. Walters CL, Arend RC, Armstrong DK, Naumann RW, Alvarez RD. Folate and folate receptor alpha antagonists mechanism of action in ovarian cancer. Gynecol Oncol. 2013;131(2):493–498.’

3. Cheung A, Bax HJ, Josephs DH, et al. Targeting folate receptor alpha for cancer treatment. Oncotarget. 2016;7(32):52553–52574.

4. Scaranti M, Cojocaru E, Banerjee S, Banerji U. Exploiting the folate receptor α in oncology. Nat Rev Clin Oncol. 2020;17(6):349–359. doi:10.1038/s41571-020-0339-5

5. Lee EK, Liu JF. Antibody-drug conjugates in gynecologic malignancies. Gynecol Oncol. 2019;153(3):694–702.

6. Zamarin D, et al; Abstract AP27: A phase 2 study of TPIV200/HUFR-1 (a multi-epitope folate receptor alpha vaccine) in combination with durvalumab in patients with platinum resistant ovarian cancer. Clin Cancer Res 15 November 2019; 25 (22_Supplement): AP27. 10.1158/1557-3265.OVCASYMP18-AP27

7. Moore KN, Oza AM, Colombo N, et al. Phase III, randomized trial of mirvetuximab soravtansine versus chemotherapy in patients with platinum-resistant ovarian cancer: primary analysis of FORWARD I. Ann Oncol. 2021;32(6):757–765.

8. Manzano A, Ocaña A. Antibody-Drug Conjugates: A Promising Novel Therapy for the Treatment of Ovarian Cancer. Cancers (Basel). 2020;12(8):2223. Published 2020 Aug 9. Doi:10.3390/cancers12082223

9. O’Malley DM, Matulonis UA, Birrer MJ, et al. Phase Ib study of mirvetuximab soravtansine, a folate receptor alpha (FRα)-targeting antibody-drug conjugate (ADC), in combination with bevacizumab in patients with platinum-resistant ovarian cancer. Gynecol Oncol. 2020;157(2):379–385.

10. ImmunoGen Presents Full Results from Positive Pivotal SORAYA Trial of Mirvetuximab Soravtansine in Ovarian Cancer at SGO Annual Meeting. BusinessWire. 19 Mar 2022. Accessed 25 Aug 2022. Online, Available <https://investor.immunogen.com/news-releases/news-release-details/immunogen-presents-full-results-positive-pivotal-soraya-trial> n.pag

11. Moore KN, Angelergues A, Konecny GE, Banerjee SN, Pignata S, Colombo N, et al. Phase III MIRASOL (GOG 3045/ENGOT-ov55) study: Initial report of mirvetuximab soravtansine vs. investigator’s choice of chemotherapy in platinum-resistant, advanced high-grade epithelial ovarian, primary peritoneal, or fallopian tube cancers with high folate receptor-alpha expression. JCO. 2023 Jun 10;41(17_suppl):LBA5507–LBA5507.

12. Naumann RW, Marin LP, Oaknin, A, et al. STRO-002-GM2: A Phase 1 Open-Label, Safety, Pharmacokinetic and Preliminary Efficacy Study of STRO-002, an Anti-Folate Receptor Alpha Antibody Drug Conjugate in Combination with Bevacizumab in Patients with Advanced Epithelial Ovarian Cancer. Abstract presented at ASCO 2022 Annual Meeting, Chicago, IL.

13. Shimizu T, Fujiwara Y, Yonemori K, et al. First-in-Human Phase 1 Study of MORAb-202, an Antibody-Drug Conjugate Comprising Farletuzumab Linked to Eribulin Mesylate, in Patients with Folate Receptor-α-Positive Advanced Solid Tumors. Clin Cancer Res. 2021;27(14):3905–3915. Doi:10.1158/1078-0432.CCR-20-4740

14. Konner JA, Bell-McGuinn KM, Sabbatini P, et al. Farletuzumab, a humanized monoclonal antibody against folate receptor alpha, in epithelial ovarian cancer: a phase I study. Clin Cancer Res. 2010;16(21):5288–5295. doi:10.1158/1078-0432.CCR-10-0700

15. Lorusso PM, Edelman MJ, Bever SL, et al. Phase I study of folate conjugate EC145 (Vintafolide) in patients with refractory solid tumors. J Clin Oncol. 2012;30(32):4011–4016. doi:10.1200/JCO.2011.41.4946

16. Naumann RW, Coleman RL, Burger RA, et al. PRECEDENT: a randomized phase II trial comparing vintafolide (EC145) and pegylated liposomal doxorubicin (PLD) in combination versus PLD alone in patients with platinum-resistant ovarian cancer. J Clin Oncol 2013;31:4400–4406.

17. Ponte JF, Ab O, Lanieri L, et al. Mirvetuximab Soravtansine (IMGN853), a Folate Receptor Alpha-Targeting Antibody-Drug Conjugate, Potentiates the Activity of Standard of Care Therapeutics in Ovarian Cancer Models. Neoplasia. 2016;18(12):775–784. Doi:10.1016/j.neo.2016.11.002

18. Data used in this publication were generated by the Clinical Proteomic Tumor Analysis Consortium (NCI/NIH)

19. Moore KN, O’Malley DM, Vergote I, et al. Safety and activity findings from a phase 1b escalation study of mirvetuximab soravtansine, a folate receptor alpha (FRα)-targeting antibody-drug conjugate (ADC), in combination with carboplatin in patients with platinum-sensitive ovarian cancer. Gynecol Oncol. 2018;151(1):46–52. Doi:10.1016/j.ygyno.2018.07.017

20. Matulonis U, Moore KN, Martin, LP, Vergote IB, Castro C, Gilbert L, Malek K, Birrer MJ, O’Malley DM. Mirvetuximab soravtansine, a folate receptor alpha (FRα)-targeting antibody-drug conjugate (ADC), with pembrolizumab in platinum-resistant ovarian cancer (PROC): Initial results of an expansion cohort from FORWARD II, a phase Ib study. Abstracts Gynaecological Cancer 10/2018: Vol 29 Supplement 8, VIII339. doi:10.1093/annonc/mdy285.157.

21. O’Malley DM, Matulonis UA, Birrer MJ, et al. Phase Ib study of mirvetuximab soravtansine, a folate receptor alpha (FRα)-targeting antibody-drug conjugate (ADC), in combination with bevacizumab in patients with platinum-resistant ovarian cancer. Gynecol Oncol. 2020;157(2):379–385. Doi:10.1016/j.ygyno.2020.01.037

22. O’Malley DM, Richardson DL, Vergote IB, Gilbert L, Castro C, Provencher D, Matulonis UA, Mantia-Smaldone G, Martin L, Zweidler-McKay PA, Moore KN. Mirvetuximab soravtansine (MIRV), a folate receptor alpha (FRα)-targeting antibody-drug conjugate (ADC), in combination with carboplatin (CARBO) and bevacizumab (BEV): Final results from a study in patients with recurrent platinum sensitive ovarian cancer. Annals of Oncology (202) 31 (supp_4):S551–S589. DOI: 10.1016/annonc/annonc276.

23. Safran M, Rosen N, Twik M, BarShir R, Iny Stein T, Dahary D, Fishilevich S, and Lancet D. *The GeneCards Suite* Chapter, Practical Guide to Life Science Databases (2022) pp 27–56

24. Karakas B, Bachman KE, Park BH. Mutation of the PIK3CA oncogene in human cancers. Br J Cancer. 2006;94(4):455–459. doi:10.1038/sj.bjc.6602970

25. Mishra R, Patel H, Alanazi S, Kilroy MK, Garrett JT. PI3K Inhibitors in Cancer: Clinical Implications and Adverse Effects. Int J Mol Sci. 2021;22(7):3464. Published 2021 Mar 27. doi:10.3390/ijms22073464

26. Konstantinopoulos PA, Barry WT, Birrer M, et al. Olaparib and α-specific PI3K inhibitor alpelisib for patients with epithelial ovarian cancer: a dose-escalation and dose-expansion phase 1b trial. Lancet Oncol. 2019;20(4):570–580. doi:10.1016/S1470-2045(18)30905-7

27. André F, Ciruelos E, Rubovszky G, et al. Alpelisib for *PIK3CA*-Mutated, Hormone Receptor-Positive Advanced Breast Cancer. N Engl J Med. 2019;380(20):1929–1940. doi:10.1056/NEJMoa1813904

28. Gasparri ML, Bardhi E, Ruscito I, et al. PI3K/AKT/mTOR Pathway in Ovarian Cancer Treatment: Are We on the Right Track? Geburtshilfe Frauenheilkd. 2017;77(10):1095–1103. doi:10.1055/s-0043-118907

29. Ruan GX, Kazlauskas A. Axl is essential for VEGF-A-dependent activation of PI3K/Akt. EMBO J. 2012;31(7):1692–1703. doi:10.1038/emboj.2012.21

30. Krook MA, Reeser JW, Ernst G, et al. Fibroblast growth factor receptors in cancer: genetic alterations, diagnostics, therapeutic targets and mechanisms of resistance. Br J Cancer. 2021;124(5):880–892. doi:10.1038/s41416-020-01157-0

31. Backes F, Fowler J, Copeland L, Wei L, O’Malley D, Cohn D, Cosgrove C, Hays J, Bixel K. Phase I Study of Mirvetuximab Soravtansine (Mirv) and Rucaparib for Recurrent Endometrial, Ovarian, Fallopian Tube or Primary Peritoneal Cancer. International Journal of Gynecological Cancer. 2021;31:A13-A4. doi: 10.1136/ijgc-2021-IGCS.24. PubMed PMID: WOS:000773625600025.

